# Disparities in Kangaroo Care for Premature Infants in the Neonatal Intensive Care Unit

**DOI:** 10.1101/2020.11.09.20224766

**Authors:** Edith Brignoni-Pérez, Melissa Scala, Heidi M. Feldman, Virginia A. Marchman, Katherine E. Travis

## Abstract

**OBJECTIVES:** The aim of this study was to investigate whether preterm infants whose families have lower socioeconomic status (SES) or communicate with clinical staff in a language other than English experience differences in the total amount, frequency, and duration of Kangaroo Care (KC) compared to preterm infants of higher SES or primarily English-speaking families.

**METHODS:** Participants were infants born <32 weeks gestational age (GA), *N*=116. We defined family SES by the infants’ health insurance (private/higher vs. public/lower) and family language by the language mothers used to communicate with clinical staff (English vs. Other language). Family SES or family language groups were compared on: (1) the total amount of KC infants experienced during hospitalization; (2) frequency of KC per visitation days; and, (3) duration of KC events per day.

**RESULTS:** Infants in the lower SES and Other language groups experienced KC in reduced amounts, lower frequencies, and shorter durations than infants in either the higher SES or English language groups. SES and language group differences remained significant after controlling for family visitation and GA at birth. After controlling for SES, language group differences in KC duration remained significant.

**CONCLUSIONS:** Our findings revealed disparities in the total amount, frequency, and duration of KC in the neonatal intensive care unit as a function of both family SES and language families used to communicate with clinical staff. These disparities reduced infants’ access to this developmental care practice shown to stabilize clinical status and promote neurodevelopment. We recommend that hospital nurseries implement policies that minimize such disparities.

**Table of Contents Summary:** Total amount, frequency, and duration of Kangaroo Care for preterm infants in the NICU varied as a function of family’s socioeconomic status and language.

**What’s Known on This Subject:** In the United States, disparities in health care delivery and medical outcomes have been identified on the basis of patient or family socioeconomic status and the language patients or families use to communicate with clinical staff.

**What This Study Adds:** Extending to the NICU, the amount, frequency, and duration of Kangaroo Care experienced by preterm infants differed both by family’s socioeconomic status and the language families use to communicate with clinical staff. Policy changes are needed to reduce these disparities.

**Contributors’ Statement Page:** Dr. Brignoni-Pérez conceptualized and designed the study, acquired data from the electronic medical record, analyzed the data, drafted the initial manuscript, and reviewed and revised the manuscript. Drs. Scala, Marchman, Feldman, and Travis conceptualized and designed the study, supervised data abstraction and analysis, and critically reviewed and revised the manuscript for important intellectual content. All authors approved the final manuscript as submitted and agree to be accountable for all aspects of the work.

## INTRODUCTION

Kangaroo Care (KC) is a developmental care practice that provides parent-to-infant skin-to-skin contact^1,2^. This practice has been shown to reduce medical complications of preterm birth, such as hypothermia, sepsis, and rehospitalization^1,2^. It has also been shown to promote infant growth, breastfeeding, and mother-infant attachment^3^. Moreover, KC has been associated with improved neurocognitive developmental outcomes, including better hearing, speech, social, and executive function skills, in preterm infants and children^4,5^. All these benefits are crucial for infants born very preterm (<32 weeks gestational age, GA), who are likely to encounter many complications of prematurity that lead to early clinical instability, prolonged hospitalization, and potentially long-term behavioral, cognitive, social-emotional, language, and learning delays^6–14^. Developmental care practices have been adopted as a part of the standard of medical care in many neonatal intensive care units (NICU)^15–19^ to reduce preterm-birth-related morbidities, support parent-infant bonding, and to possibly improve long-term developmental outcomes. Despite the many apparent benefits of KC, however, several barriers may reduce opportunities for such practice, including parental factors (e.g., rates of visitation, family comfort with the practice) and health system factors (e.g., unit design, adequate staff support, parent educational programs, access to translators)^3,20–28^. Parents and health care providers are also susceptible to cultural norms and personal beliefs that affect the frequency and amount of KC^20,21,25^.

Studies have shown that child and adult patients with lower socioeconomic status (SES) are consistently less healthy than wealthier and more educated patients^29^. Differences in health care delivery and medical practices contribute to these disparities. Such findings extend to the NICUs in the United States, where families experience disparities in health care on the basis of socioeconomic and cultural/ethnic factors^30^. It has been shown that NICUs with higher proportions of patients from lower SES have overall lower quality care as measured by a composite of maternal and infant outcome measures^31^. Additionally, health insurance (private vs. public), a proxy of SES, relates to prenatal and postnatal health opportunities, negatively impacting the health of those with lower SES^32^. Moreover, in the United States, non-English-speaking families in the NICU have been shown to be more susceptible to misunderstanding their child’s diagnosis and treatments because clinical staff and families do not share a common language^33^. To date, there is limited information about whether and how parents’ opportunities to provide KC for their prematurely born infant in the NICU are influenced by their socioeconomic and linguistic background^3^, despite the beneficial role of this developmental care practice. In the present study, we investigated whether the family’s SES and their preferred language to communicate with clinical staff influenced the total amount, frequency, and duration of KC with their preterm infants in the NICU. We hypothesized that infants whose families have lower SES or communicate with clinical staff in a language other than English would receive lower total amount, frequency, and duration of KC compared to infants whose families have higher SES or speak English to clinical staff. These findings would have implications for building policies and procedures to increase KC in the NICU for groups at increased risk for adverse health and developmental outcomes.

## METHODS

Participants were infants born at a GA of less than 32 weeks, who were hospitalized at the Lucile Packard Children Hospital (LPCH) in Stanford. From the electronic medical record (EMR), we retrospectively acquired data on these infants’ experience of KC from May 1, 2018, when developmental care practices (including KC) at LPCH started being recorded consistently in the EMR by clinical staff, to March 8^th^, 2020, when LPCH changed visitation policies due to COVID-19. These infants’ data are part of a broader study investigating developmental care practices in relation to brain development, and thus we collected the data from the date of birth (DOB) until the infants’ routine brain imaging session date (MRI) that occurs around 36 weeks postmenstrual age.

The sample (*N*=116) was divided into two groups by each of the two key factors: family SES and family language. For SES, we used the infant’s health insurance as a proxy for this factor. Private insurance was classified as higher SES and public insurance as lower SES (Table 1). For family language, we used a specific field in the EMR that indicated the language that mothers used to communicate with clinical staff, specifically either English or another language (e.g., Spanish, Mandarin, Dari), as a proxy for language primarily used by all family members with clinical staff. For one participant, these data were missing (i.e., the language their mother used to communicate with clinical staff), and thus we used the language reported for the father. At LPCH, translators were available at all times for most common languages either via bedside iPads or in-person interpreters during daytime hours. Of those families who used a language other than English (*n*=34), 26 families used Spanish, the largest linguistic representation in the study location that is not English. The protocols for this study were approved by the Stanford University Institutional Review Board.

**Table 1:** Description of the participants by family socioeconomic status.

|  | *Higher*<br>*n (%) or M (SD)* | *Lower*<br>*n (%) or M (SD)* | *X<sup>2</sup> or t* | *p* |
| --- | --- | --- | --- | --- |
| **Demographics** |  |  |  |  |
| Total *N* | 62 | 54 |  |  |
| Sex: Female | 27 (44) | 24 (44) | 0.01 | 0.92 |
| GA at Birth (weeks) | 28.5 (2.5) | 29.2 (2.3) | -1.57 | 0.12 |
| Hospitalization Duration (days) | 62 (32) | 53 (27) | 1.53 | 0.13 |
| Family Language Use: English | 55 (89) | 27 (50) | **20.87** | **<0.001** |
| Race: White | 13 (21) | 10 (19) | 0.11 | 0.74 |
| **Clinical Factors** |  |  |  |  |
| Apgar 1 (minutes) | 5 (2) | 6 (2) | -0.56 | 0.58 |
| Apgar 5 (minutes) | 7 (1) | 8 (2) | -0.76 | 0.45 |
| Necrotizing Enterocolitis | 5 (8) | 5 (9) | 0.05 | 0.82 |
| High Frequency Oscillatory Ventilator | 8 (13) | 6 (11) | 0.13 | 0.72 |
| **Family Visitation** |  |  |  |  |
| Visitation Days/Hospital Days (%) | 77 (18) | 65 (24) | **2.94** | **<0.001** |

*Abbreviations:* GA = gestational age; M = mean; SD = standard deviation; bold = significant.

### Population description, clinical risk, and family visitation

We extracted the following information about the participants to characterize the sample: sex, GA at birth, days of hospitalization (from DOB to MRI), and race. We also extracted information about infants’ medical conditions, including clinical factors that may have reduced infants’ ability to receive KC or barriers to KC per policy in the LPCH NICU (e.g., necrotizing enterocolitis, proportion of infants requiring high frequency oscillatory ventilator). To account for parents’ availability to perform KC, we determined the frequency of family visitation from the EMR. At LPCH, parents were permitted to visit at any time of the day, except during nursing sign out (7:00-7:30 a.m./p.m.). Daily visitation was counted as having occurred if clinical staff charted that any family member engaged in KC with their infant or had visited at bedside. We quantified the frequency of family visitation as the percentage of days that families visited out of the total days an infant was hospitalized (from DOB to MRI).

### Kangaroo Care

The NICU at LPCH has standardized unit guidelines on developmental care activities to support parent participation. KC done by any family member was recorded by clinical staff in the infants’ EMR. We derived the following three metrics from the KC data to assess: (1) KC total amount, the total minutes infants experienced KC from DOB to MRI; (2) KC frequency, the percentage of days that families performed KC out of the total number of days that families visited the hospital (total KC days/total visitation days); and, (3) KC duration, the rate in minutes per day of KC events (total KC minutes/total KC days).

### Data analyses

We performed separate Chi square tests for each categorical variable and independent samples t-tests for each continuous variable to compare groups on demographic, clinical, and visitation variables. For KC total amount, frequency, and duration, we performed separate analyses by each group factor: family SES or family language. Since the metric KC total amount was intrinsically biased by the variance in visitation days across infants, we performed a univariate analysis of covariance (ANCOVA), controlling for family visitation. We also performed separate univariate ANCOVA to control for demographic or clinical risk factors that were found to differ between the groups. For KC frequency and duration metrics, we carried out separate analysis of variance by family SES or family language. Also, we performed separate univariate ANCOVA to control for each demographic or clinical risk factor that differed between the groups. Threshold for significance was set at *p* < 0.05. All analyses were conducted using the Statistical Package for Social Sciences 26 (https://www.ibm.com/support/pages/downloading-ibm-spss-statistics-26).

## RESULTS

### Population description, clinical factors, and family visitation

Table 1 shows demographic, clinical, and family visitation variables of the sample divided by family SES. The groups were balanced in all demographic variables but family language. The lower SES group had a higher proportion of infants whose families used a language other than English to communicate with clinical staff. The groups did not statistically differ on clinical factors. The percentage of days infants in the lower SES group were visited by their families was significantly lower compared to infants in the higher SES group.

Table 2 shows demographic, clinical, and family visitation variables of the sample divided by family language. The groups were statistically balanced on all demographic variables other than GA at birth and family SES; the Other language group had infants born at an older GA and were predominantly from lower SES families. The groups did not statistically differ on clinical factors. The percentage of days infants in the Other language group were visited by their families was lower, although not statistically significant, relative to infants in the English group.

**Table 2:** Description of the participants by family language use.

|  | *English*<br>*n (%) or M (SD)* | *Other Language*<br>*n (%) or M (SD)* | *X<sup>2</sup> or t* | *p* |
| --- | --- | --- | --- | --- |
| **Demographics** |  |  |  |  |
| Total *N* | 82 | 34 |  |  |
| Sex: Female | 34 (41) | 17 (50) | 0.71 | 0.40 |
| GA at Birth (weeks) | 28.5 (2.5) | 29.7 (2.0) | **-2.63** | **0.01** |
| Hospitalization Duration (days) | 61 (30) | 52 (31) | 1.40 | 0.16 |
| Family Socioeconomic Status: Higher | 55 (67) | 7 (21) | **20.87** | **<0.001** |
| Race: White | 20 (24) | 3 (9) | 3.66 | 0.06 |
| **Clinical Factors** |  |  |  |  |
| Apgar 1 (minutes) | 5 (2) | 6 (2) | -1.30 | 0.20 |
| Apgar 5 (minutes) | 7 (1) | 8 (2) | -1.40 | 0.16 |
| Necrotizing Enterocolitis | 9 (11) | 1 (3) | 1.97 | 0.16 |
| High Frequency Oscillatory Ventilator | 12 (15) | 2 (6) | 1.84 | 0.17 |
| **Family Visitation** |  |  |  |  |
| Visitation Days/Hospital Days (%) | 74 (21) | 66 (23) | 1.89 | 0.06 |

*Abbreviations:* GA = gestational age; M = mean; SD = standard deviation; bold = significant.

### Kangaroo Care

Table 3 compares KC metrics between family SES groups. The total amount, frequency, and duration of KC was significantly lower for infants in the lower SES group, as compared to the higher SES group (Table 3; Figure 1 A-C). Between-group differences in the total amount of KC remained significant after controlling for family visitation and either GA at birth or family language. Group differences in frequency and duration of KC also remained significant after controlling for GA at birth or family language.

**Figure 1.**
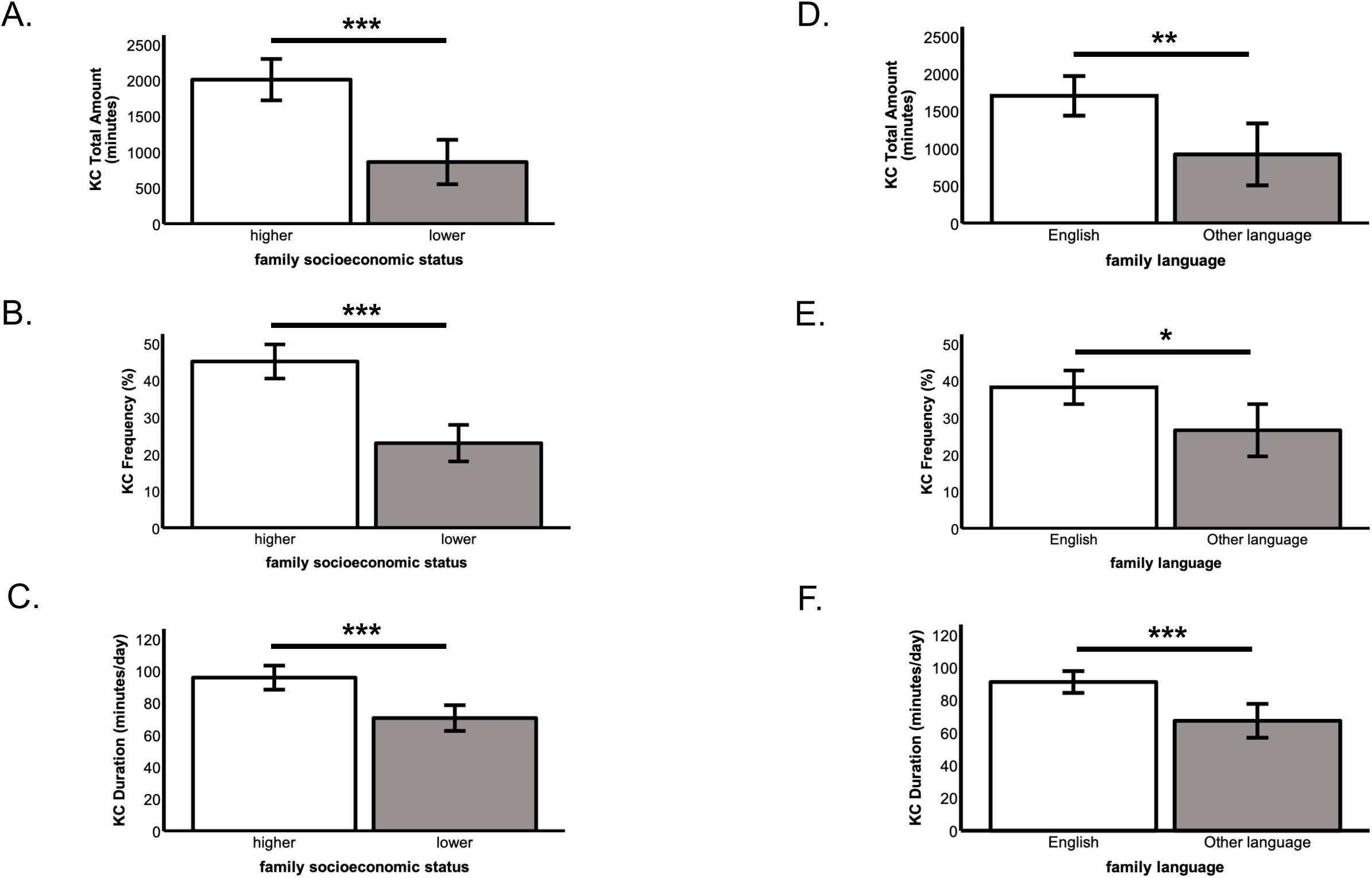
Results of Kangaroo Care by family SES (A-C) or family language (D-F). **A**. Total amount of KC during hospitalization by family SES, controlling for family visitation. **B**. Frequency of KC days out of family visitation days by family SES. **C**. Duration of KC events during KC days by family SES. **D**. Total amount of KC during hospitalization by family language, controlling for family visitation. **E**. Frequency of KC days out of family visitation days by family language. **F**. Duration of KC events during KC days by family language. * = *p* < 0.05; ** = *p* < 0.01; *** = *p* < 0.001. Error bars = standard error of the mean.

**Table 3:**
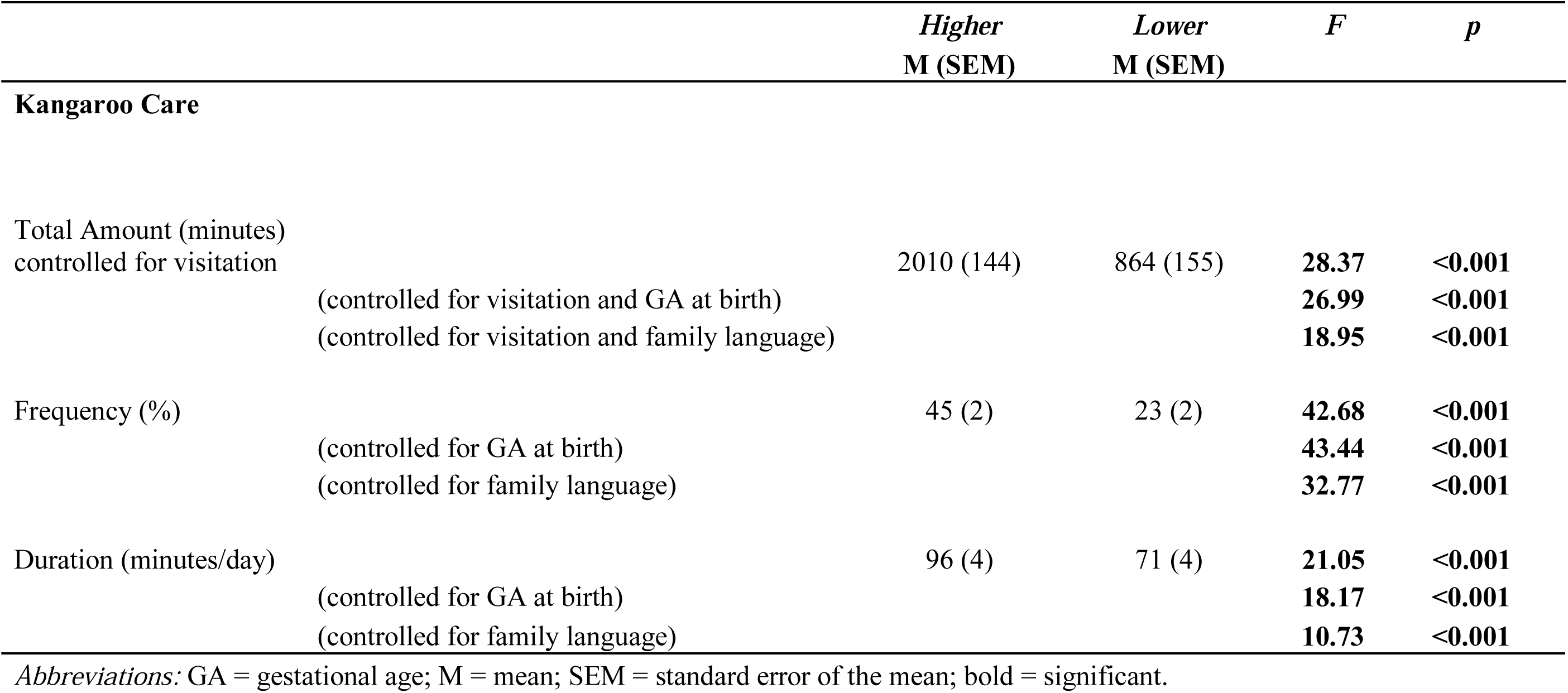
Results of Kangaroo Care by family socioeconomic status.

Table 4 compares KC metrics between family language groups. The total amount, frequency, and duration of KC was significantly lower for infants in the Other language, as compared to the English language group (Table 4; Figure 1 D-F). Between-group differences in the total amount of KC remained significant after controlling for family visitation and GA at birth, but not when controlling for family visitation and family SES. Group differences in frequency of KC remained significant after controlling for GA at birth, but not family SES. Group differences in the duration of KC remained significant after controlling for either GA at birth or family SES.

**Table 4:** Results of Kangaroo Care by family language use.

|  | *English*<br>M (SEM) | *Other Language*<br>M (SEM) | *F* | *p* |
| --- | --- | --- | --- | --- |
| **Kangaroo Care** |  |  |  |  |
| Total Amount (minutes)<br>controlled for visitation | 1706 (133) | 923 (207) | **10.00** | **<0.001** |
| (controlled for visitation and GA at birth) |  |  | **5.04** | **0.03** |
| (controlled for visitation and SES) |  |  | 1.93 | 0.17 |
| Frequency (%) | 38 (2) | 27 (3) | **7.63** | **0.01** |
| (controlled for GA at birth) |  |  | **7.99** | **0.01** |
| (controlled for SES) |  |  | 0.16 | 0.69 |
| Duration (minutes/day) | 91 (3) | 67 (5) | **14.83** | **<0.001** |
| (controlled for GA at birth) |  |  | **10.35** | **<0.001** |
| (controlled for SES) |  |  | **5.03** | **0.03** |

*Abbreviations:* GA = gestational age; SES = socioeconomic status; M = mean; SEM = standard error of the mean; bold = significant.

## DISCUSSION

Consistent with our hypothesis, we found that KC, however measured, total amount during hospitalization, frequency during visitation, and duration per day, was experienced less by infants from lower SES families or whose families spoke a language other than English. These results were significant after controlling for days of family visitation and for infants’ GA at birth. Disparities in KC found on the basis of family SES also remained significant after controlling for family language. Disparities found on the basis of family language for KC duration, but not total amount or frequency, remained significant after controlling for family SES.

The present study provides novel evidence that both family SES and family language other than English are factors that contribute to significant disparities in the total amount, frequency, and duration of KC experienced by preterm infants in the NICU. Importantly, we show that disparities in KC were not solely explained by how frequently families visited the hospital. These findings are generally consistent with previous studies that have examined the contribution of socio-demographic factors to parental involvement in KC activities. For example, a study reported that parental holding increases if mothers are White, married, older, or employed compared to non-White, single, and younger parents ^34^. Another study showed that white mothers are told KC is an activity they could do with their infants 50% more often compared to non-white mothers^35^. Finally, a survey of parents at multiple European NICUs found longer periods of KC are associated with increased maternal education^36^. More research is needed to identify specific barriers in subgroups of disadvantaged populations in the NICU on the basis of both family SES and family language.

KC is reported to improve many clinical^1–3^ and neurocognitive developmental^4,5^ outcomes. The optimum dose and timing of KC needed to confer these advantages is unknown. However, periods of at least 60 minutes per day are generally recommended. Longer periods may better support infant sleep cycles and offer increased benefits in infant growth, cardiorespiratory stability, and neurodevelopment^37,38^. To date, the risks of too much KC have seemed small; thus, general policies have been to encourage as much as an infant can tolerate medically. If specific thresholds of medical benefit exist, the infants of lower SES families and non-English-speaking families in the present study may be at a further disadvantage in reaching goal doses of this important developmental care practice, therefore raising further concern for the outcomes of these already at-risk infants.

The present findings reveal that both family SES and family language should be individual targets for programmatic interventions to reduce disparities in KC. Recent studies have shown that both disadvantaged individual (e.g., psychological and physical wellness) and systemic (e.g., parental leave policies) factors impede the frequency and duration of KC provided by parents in the NICU^25,26^. Barriers to KC may also overlap with those known to impede parental visitation, such as unemployment or low income, lack of support for other children (e.g., childcare), health insurance, marital status, parents’ age, transportation, and work or household responsibilities^39,40^. Those barriers that can be addressed at the system-level should include, for example, childcare in the hospital for other children in the family, transportation to/from the home^25,41^, and increased training of clinical staff on interpreter use^42^. Of note, the NICU at LPCH has fairly robust resources compared to many other locations in the United States and yet disparities persist. Until now, most evaluations of disparities in NICU care were done using larger databases or multicenter qualitative studies^30,43,44^. While these studies provide certain insights, they can inadvertently lead centers to conclude that the problem exists elsewhere. The single-center construct of our study is limited by sample size but represents a potentially powerful example of critical self-reflection necessary for actionable change through quality improvement initiatives or further interventional research.

It is also possible that the role of family SES and family language may operate more indirectly to impact care delivery. For example, these factors may impact the quality of interaction between parents and clinical staff. Rates of parent visitation have been shown to be significantly correlated with parental stress and communication with clinical staff^45^ and to improve when programs are implemented to provide individualized encouragement for maternal visitation^46^. Possible strategies are policies that promote family-centered approaches in which parents are seen as partners of the clinical staff^47,48^ and which expect parents to be present for longer periods and transition to active caregivers, thus potentially removing many barriers to visitation and to KC^49^. In societies without social supports of extended parental leave and childcare programs, these offered models are likely to have little impact on disparities between groups^50^. More research is needed to understand the potential impact of family-centered care programs on mitigating disparities, particularly in countries with fewer social programs to support parents.

Another indirect factor could be ambivalence among NICU clinical staff regarding the importance of family involvement and KC in NICU care, in spite of evidence for its benefits and endorsement by national organizations. In a 2013 survey of NICU parents and nurses, 63% of mothers but only 18% of nurses felt that KC should be offered daily, and 90% of mothers compared to 40% of nurses felt mothers should be partners in care^51^. While parents may not understand medical barriers to KC, disagreements or communication barriers between family and clinical staff only hamper efforts at family-centered care. Most national quality metrics used to gauge NICU care do not include direct measures of family-centered care; best family-centered care measures are still under discussion^52^. Clinical staff may interpret quality standards as stressing factors, such as equipment dislodgements like unplanned extubations, as more important than parent engagement in infant care. National quality standards should include measures of family-centered developmental care, as well as direct measures of disparate care.

Peer support might be an additional support to clinical staff that may help to reduce disparities in KC. Social support is a key component of health care; thus, training families in the NICU to support their peers (e.g., language use/communication, encouragement) could contribute to the family-centered approach to improve access to this important experience^53,54^. It has also been argued that nurses have the ability to ameliorate many of the barriers that parents encounter to participate as partners in their premature infants’ health care, given their important role in the NICU^48^.

Overall, the current findings emphasize the critical issue of equity. The observed disparities in KC in relation to the family’s socioeconomic and language background represent a challenge to clinical staff in the NICU. Health care professionals must address the need to provide families of lower SES and those who speak a language other than English with the resources and services they need to provide comparable opportunity as enjoyed by infants from wealthier backgrounds and primarily English-speaking families. To achieve equity, NICUs may need to write or modify policies and practices for increasing parents’ visitation, then increasing family education and support when they are at the hospital, and finally addressing medical and nursing practices and education to support families to begin and sustain KC. In addition, a quality improvement approach, now required in many medical settings and in training, may encourage rapid change faster than could be achieved with intervention studies. Quality improvement would allow an iterative process based on intervention and careful measurement^55–57^, leading to reduced disparities and ultimately improved outcomes for all infants.

This study has limitations. The data were extracted and analyzed from EMRs and may thus capture inconsistencies in reporting from clinical staff. The sample was not large enough to explore potential interactions between family SES and family language use. Our measures of family SES and family language were limited. Finally, this investigation was a single-site study. More studies are needed to further assess the barriers to KC in the NICU, specifically focused on parents-clinical staff partnership quality.

## CONCLUSIONS

This investigation presents an in-depth analysis of KC for preterm infants in a NICU in the United States in relation to their family’s SES and language use in the hospital, revealing significant and concerning disparities. We recommend rapid modifications of policies that guide and promote this developmental care practice in the NICU and quality improvement studies to assure rapid and effective quantitative changes. A common goal should be to reduce disparities in KC, a critical early-life experience in this at-risk population.

## Data Availability

De-identified data are available upon request to the senior author.

## ACKNOWLEDGMENTS

We want to thank Maya C. Morales for her assistance. This work was possible also thanks to the Stanford REDCap platform (http://redcap.stanford.edu), which is developed and operated by Stanford Medicine Research IT team. The REDCap platform services at Stanford are subsidized by a) Stanford School of Medicine Research Office and b) the National Center for Research Resources and the National Center for Advancing Translational Sciences, National Institutes of Health, through grants KL2 TR001085 and TL1 TR001085.

Also, this research used data or services provided by STARR, “STAnford medicine Research data Repository”, a clinical data warehouse containing live EMR data from Stanford Health Care, the Stanford Children’s Hospital, the University Healthcare Alliance and Packard Children’s Health Alliance clinics, and other auxiliary data from Hospital applications, such as radiology PACS. STARR platform is developed and operated by Stanford Medicine Research IT team and is made possible by Stanford School of Medicine Research Office.

## Abbreviations

ANCOVA: analysis of covariance
DOB: date of birth
EMR: electronic medical record
GA: gestational age
KC: Kangaroo Care
LPCH: Lucile Packard Children’s Hospital
MRI: magnetic resonance imaging
NICU: neonatal intensive care unit
SES: socioeconomic status

## Notes

**Funding/Support:** This research work was supported by grants from the *Eunice Kennedy Shriver* National Institute of Child Health and Human Development (K.E. Travis, PI: 5R00HD8474904; H.M. Feldman, PI: 2RO1- HD069150) and the National Institute of Mental Health Postdoctoral Research Training in Child Psychiatry and Neurodevelopment (A. Reiss, PI: T32 MH019908).

### Competing Interest Statement

The authors have declared no competing interest.

### Funding Statement

This research work was supported by grants from the Eunice Kennedy Shriver National Institute of Child Health and Human Development (K.E. Travis, PI: 5R00HD8474904; H.M. Feldman, PI: 2RO1- HD069150) and the National Institute of Mental Health Postdoctoral Research Training in Child Psychiatry and Neurodevelopment (A. Reiss, PI: T32 MH019908).

### Author Declarations

The protocols for this study were approved by the Stanford University Institutional Review Board.

